# Protective lipid-lowering genetic variants in healthy older individuals without coronary heart disease

**DOI:** 10.1101/2021.02.16.21251811

**Authors:** Paul Lacaze, Moeen Riaz, Robert Sebra, Amanda J Hooper, Jing Pang, Jane Tiller, Galina Polekhina, Andrew M Tonkin, Christopher M Reid, Sophia Zoungas, Anne M Murray, Stephen J Nicholls, Gerald F Watts, Eric Schadt, John J McNeil

## Abstract

**Background:** Disruptive genetic variants in the *PCSK9* and *APOB* genes result in lower serum low-density lipoprotein cholesterol (LDL-C) levels and confer protection against coronary heart disease (CHD). Few studies have measured the prevalence and selective advantage of such variants among healthy older individuals without prior CHD events.

**Methods and Results:** We performed targeted sequencing of the *PCSK9* and *APOB* genes in 13,131 healthy older individuals without CHD aged 70 years or older enrolled into the ASPirin in Reducing Events in the Elderly (ASPREE) trial. We detected predicted loss-of-function (pLoF) variants in the *PCSK9* and *APOB* genes, and associated variant carrier status with blood lipid levels. We detected 22 different rare *PCSK9/APOB* candidate variants with lipid-lowering effect, carried by 104 participants (carrier rate 1 in 126). Rare variant carrier status was associated with 19.4 mg/dl (14.6%) lower LDL-C, compared with non-carriers (P=<0.001, adjusted for statin use). Statin prescriptions were less prevalent in rare variant carriers (16%) than non-carriers (35%). The *PCSK9* R46L variant (rs11591147-T) was associated with 15.5mg/dl (11.8%) lower LDL-C in heterozygotes, and 25.2 mg/dl (19.2%) lower LDL-C in homozygotes, respectively (both P=<0.001).

**Conclusions:** Lipid-lowering genetic variants are carried by healthy older individuals and contribute to CHD-free survival.

## INTRODUCTION

Genetic variants that lower serum low-density lipoprotein cholesterol (LDL-C) levels have been demonstrated to be protective against coronary heart disease (CHD) ^1-9^. In particular, protection can be conferred by rare loss-of-function, protein-truncating variants in canonical lipid-metabolism genes, including the apolipoprotein B (*APOB*) ^3^ and proprotein convertase subtilisin kexin type 9 (*PCSK9*) ^3,10^ genes. These protective genetic variants tend to be rare in the population. In addition, LDL-C particle size and other lipoprotein-related genotypes have been associated with CHD-free longevity ^11^.

Familial hypobetalipoproteinemia is caused by heterozygosity for *APOB* variants that generally result in LDL-C concentrations that are >50% lower than normal, while *PCSK9* loss-of-function variants are associated with more modest effects of 15-40% lower LDL-C ^12^. Discovery and understanding of rare protective variants in these genes, particularly *PCSK9*, has informed the successful development of several lipid-lowering therapies ^13^. However, most cholesterol-lowering variants, to date, have been identified from case-control or population-based studies ^1-9^. Healthy older populations without CHD represent an under-studied resource for the discovery and understanding of protective lipid-modifying genetic variants that may reduce CHD risk.

With this rationale, we sequenced 13,131 healthy older individuals without CHD enrolled in the ASPirin in Reducing Events in the Elderly (ASPREE) trial ^14^, with average age 75 years. Among this healthy older CHD-free population, we measured the prevalence of rare cholesterol-lowering variants in canonical lipid metabolism genes *PCSK9* and *APOB*, and associated variant carrier status with serum LDL-C and total cholesterol (TC) levels.

## METHODS

### Study population

Participants were enrolled in the ASPREE study, a randomized, placebo-controlled trial of daily low-dose aspirin ^15-17^. The ASPREE study design ^18,19^, recruitment ^20^, and baseline characteristics ^14^ have been published previously. Participants had no previous diagnosis of atherosclerotic or atherothrombotic cardiovascular disease, including myocardial infarction; heart failure; angina; pectoris; stroke, or transient ischemic attack; or diagnosis of atrial fibrillation or high blood pressure ^21^. Diagnosis of dementia or other serious illness likely to cause death within 5 years were also exclusion criteria. Genetic analysis was conducted on 13,131 samples provided by Australian ASPREE participants aged 70 years or older at enrolment ^22^. Ethics approval for genetic analysis was obtained from the Alfred Hospital Human Research Ethics Committee.

### DNA sequencing and variant analysis

A targeted sequencing panel was designed containing the *PCSK9* and *APOB* genes ^22^. Following standard protocols, DNA was extracted and sequenced using the Thermo Fisher Scientific S5TM XL system to average 200X depth, with sequences aligned to the human genome reference 37. We identified candidate cholesterol-lowering variants from sequence data using two methods: 1) prediction of rare candidate loss-of-function, protein-truncating *PCSK9* and *APOB* variants using the Loss-Of-Function Transcript Effect Estimator (LoFTEE) tool, high-confidence filter, a plugin of Ensemble Variant Effect Predictor (VEP) ^23^; and 2) assessment of candidate variants associated with hypercholesterolaemia or hypobetalipoproteinaemia identified from published functional and population studies ^3,10,24^ (for list of variants meeting the inclusion criteria, see Supplementary Materials). We also examined the effect of the *PCSK9* R46L variant with known lipid-lowering effect (rs11591147-T) ^25^. Variants were curated manually by two or more laboratory scientists following ACMG/AMP Standards ^26^ and variant classification was agnostic to lipid effects. A random selection of 10% of variants detected were validated using Sanger sequencing with 100% concordance.

### Association of variant carrier status with blood lipid levels

We sought to determine whether *PCSK9/APOB* rare variant carriers in ASPREE were associated with lower serum cholesterol levels at time of enrolment, versus age- and gender-matched non-carriers. To do this, we compared serum LDL-C and total cholesterol (TC) levels between variant carriers and N=9,540 age- and gender-matched non-carrier ASPREE controls who did not carry any *PCSK9* or *APOB* variants that met our inclusion criteria. Baseline LDL-C and TC levels were measured in routine blood samples provided by ASPREE participants at enrolment, analysed at commercial pathology laboratories. We tested association of variant carrier status with serum LDL-C and TC levels using multivariable linear regression, adjusting for age, gender, diabetes, hypertension, smoking status, alcohol use, and body mass index. We first tested association using raw unadjusted LDL-C and TC levels (not accounting for statin use), then separately using statin-adjusted levels, dividing by LDL-C and TC levels by 0.7 and 0.8 respectively for those using statin medication to estimate untreated levels, as done previously ^3^. We identified statin users based on concomitant medication data collected by ASPREE (Anatomical Therapeutic Chemical [ATC] code = C10, lipid modifying agents).

## RESULTS

Characteristics of the 13,131 sequenced participants are shown in Table 1. The median age at enrolment was 75 years; with 54% of participants female; 28% obese; and 4% current smokers. Participants had no previous diagnosis of cardiovascular disease or dementia ^14^. Most participants were of European ancestry (99% self-reported as white/Caucasian).

**Table 1.** Characteristics of Sequenced Participants at Enrolment. Sequenced participants were enrolled in the ASPREE clinical trial, aged 70 years and older (average age 75 years), without a previous diagnosis of cardiovascular disease, dementia, permanent physical disability, or current diagnosis of life-threating cancer at enrolment. Most were white/Caucasian and 54% were female.

|  | **ASPREE**<br>**N=13,131**<br>(mean age 75 years) |
| --- | --- |
| **Female sex** – no. (%) | 7056 (54) |
| **Age in years** – no. (%) |  |
| 70-74 yr | 7,894 (60) |
| 75-79 yr | 3,406 (26) |
| ≥80 yr | 1,831 (14) |
| **Race or ethnic group**<sup>a</sup> – no. (%) |  |
| White/Caucasian | 12,953 (99) |
| Other | 178 (1) |
| Obese (BMI ≥ 30 kg/m<sup>2</sup>)<sup>b</sup> (%) | 28 |
| Current smoking (%) | 4 |
| Statin use<sup>c</sup> (%) | 33 |
| Heart, stroke or vascular disease<sup>d</sup> (%) | 0 |

<sup>a</sup> Self-report.
<sup>b</sup> Obese was defined as body-mass index (weight in kilograms divided by the square of the height in meters) of ≥30.
<sup>c</sup> Anatomical Therapeutic Chemical (ATC) code C10 (lipid modifying agents).
<sup>d</sup> Baseline Characteristics of Participants in the ASPREE (ASpirin in Reducing Events in the Elderly) Study<sup>14</sup>

Among this population, we detected a total of 104 ASPREE participants carrying rare candidate loss-of-function (LoF) variants that met our inclusion criteria (MAF<0.01). This corresponded to a carrier rate of 1 in 126 participants. We detected a total of 22 different rare variants, ranging in frequency between MAF=0.00335 (detected in 44 ASPREE participants) to MAF=0.00008 (singletons detected in only one ASPREE participant) (Table 2). We found six putatively novel rare *APOB* variants that, at the time of our analysis, were not found in the gnomAD or dbSNP databases, and were not previously reported in the literature. Each of these rare *APOB* variants were detected as singletons in the ASPREE cohort (MAF=0.0008) and met our criteria for loss-of-function (Table 2).

**Table 2:**
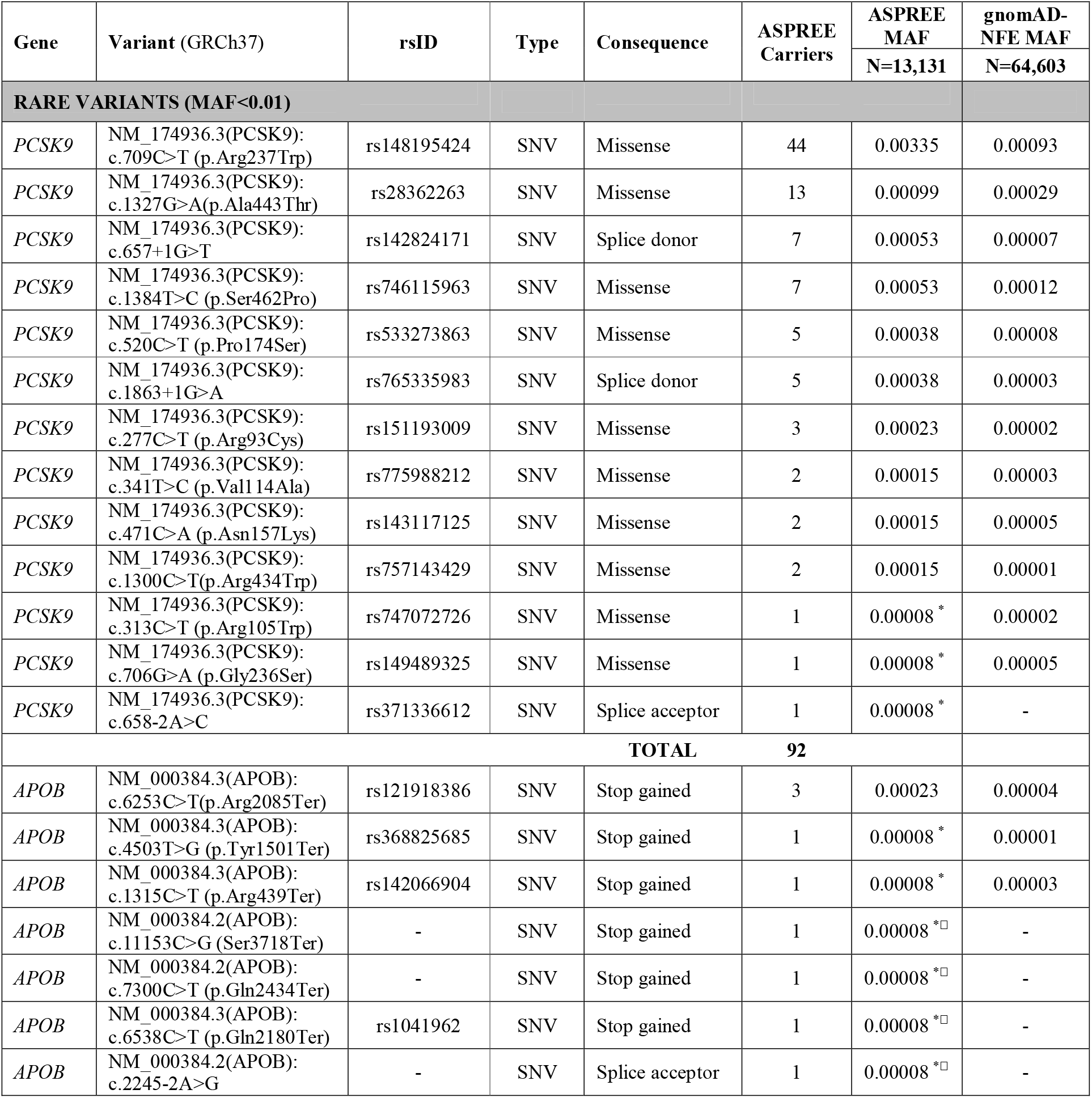
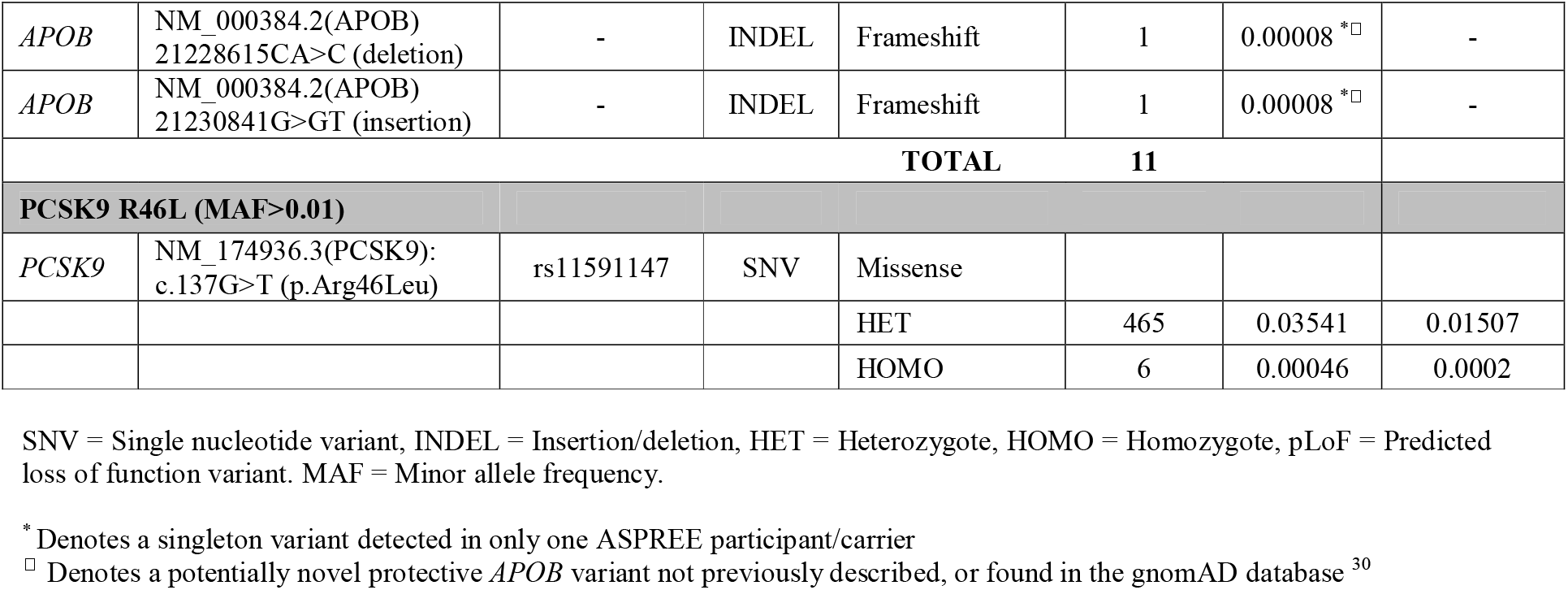
Prevalence of LDL cholesterol-lowering variants in a healthy elderly CHD-free population (ASPREE). We sequenced N=13,131 individuals aged 70 years and older to detect cholesterol-lowering variants using two methods: 1) prediction of rare candidate loss-of-function, protein-truncating *PCSK9* and *APOB* variants; and 2) assessment of candidate variants identified in previously published functional or population studies. We also examined the effect of the common *PCSK9* R46L variant with lipid-lowering effect; rs11591147 ^25^. Variants were curated following ACMG/AMP Standards ^26^.

Based on raw LDL-C and TC levels uncorrected for statin use, rare variant heterozygous carrier status in ASPREE (N=104 participants) was associated with 11.6 mg/dl (9.7%) lower serum LDL-C (P<0.001) and 7.8 mg/dl (3.9%) lower serum TC (P<0.001), versus non-carriers, adjusted for age, gender, diabetes, hypertension, smoking status, alcohol use, and BMI (Table 3). After adjusting for statin use (dividing LDL-C and TC levels by 0.7 and 0.8 respectively for those taking statin medication), rare variant heterozygous carrier status was associated with 19.4 mg/dl (14.6%) lower adjusted serum LDL-C (P<0.001) and 16.4mg/dl (7.5%) lower adjusted serum TC (P<0.001), versus non-carriers. The prevalence of lipid-lowering statin prescriptions among rare variant carriers was 16% (N=17/104), less than half that observed in non-carriers (35%, N=3324/9540).

**Table 3.** Variant carrier status in ASPREE is associated with reduced serum low-density lipoprotein cholesterol (LDL-C) and total cholesterol (TC) levels. We compared serum low-density lipoprotein cholesterol (LDL-C) and total cholesterol (TC) levels between variant carriers and N=9,540 non-carrier ASPREE controls. The multivariable linear regression model adjusted for age, gender, diabetes, hypertension, smoking status, alcohol use, and body mass index. We first tested association using raw unadjusted LDL-C and TC levels (not accounting for statin use), then separately using statin-adjusted levels (*adj*LDL-C, *adj*TC), dividing LDL-C and TC levels by 0.7 and 0.8 respectively for those using statin medication to estimate untreated levels, as done previously ^3^. We identified statin users based on concomitant medications recorded by ASPREE at baseline. Data are presented as median (inter-quartile range). Statistical significance is denoted by P<0.001. To convert values from mg/dl to mmol/l multiply by 0.02586. ATC code C10 = Anatomical Therapeutic Chemical code C10 (lipid modifying agents).

| **Rare variant group**<br>*PCSK9/APOB*<br>MAF<0.01 | **Non-Carriers**<br>N=9,540 | **Heterozygotes**<br>N=104 | **Homozygotes**<br>N=0 | **P-Value** |
| --- | --- | --- | --- | --- |
| LDL-C (mg/dl) | 119.9<br>(96.7 to 143.1) | 108.3<br>(81.2 to 131.5) | - | <0.001 |
| *adj*LDL-C (mg/dl) | 131.5<br>(112.1 to 150.8) | 112.1<br>(92.8 to 139.2) | - | <0.001 |
| TC (mg/dl) | 201.1<br>(177.9 to 228.2) | 193.3<br>(170.1 to 220.4) | - | <0.001 |
| *adj*TC (mg/dl) | 217.5<br>(193.4 to 241.6) | 201.1<br>(177.9 to 225.7) | - | <0.001 |
| **Common variant**<br>*PCSK9* rs11591147-T | **Non-Carriers**<br>N=9,540 | **Heterozygotes**<br>N=465 | **Homozygotes**<br>N=6 | **P-Value** |
| LDL-C (mg/dl) | 119.9<br>(96.7 to 143.1) | 112.1<br>(88.9 to 131.5) | 106.35<br>(95.7 to 117.0) | <0.001 |
| *adj*LDL-C (mg/dl) | 131.5<br>(112.1 to 150.8) | 116.0<br>(96.7 to 135.3) | 106.4<br>(95.7 to 117.0) | <0.001 |
| TC (mg/dl) | 201.1<br>(177.9 to 228.2) | 197.2<br>(170.1 to 216.6) | 197.2<br>(168.2 to 211.7) | <0.001 |
| *adj*TC (mg/dl) | 217.5<br>(193.4 to 241.6) | 201.1<br>(183.6 to 227.1) | 197.2<br>(168.2 to 211.7) | <0.001 |

The estimated median untreated LDL-C level in non-carriers was 131.5 mg/dl (110.4 to 154.7), compared with 112.1 mg/dl (92.8 to 139.2) in rare *PCSK9/APOB* variant carriers. At the per-gene level, rare variant carrier status for *PCSK9* and *APOB* variants separately was also associated with significantly lower LDL-C levels for both genes (P<0.001) (Figure 1).

**Figure 1.**
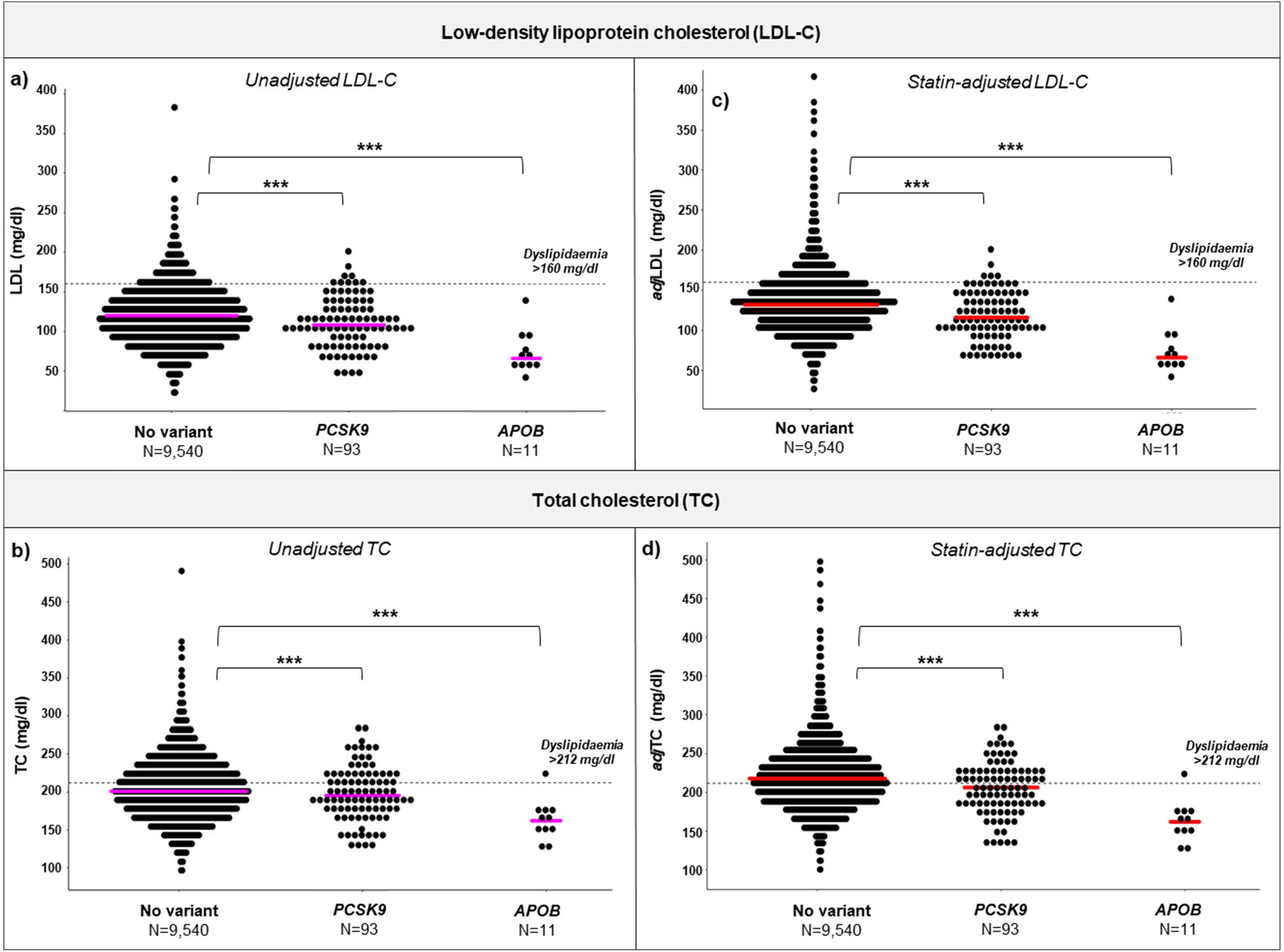
*PCSK9* and *APOB* rare variant carrier status in ASPREE is associated with reduced serum lipoprotein cholesterol (LDL-C) and total cholesterol (TC) levels. Shown is a comparison of LDL-C and TC levels at enrolment for carriers of rare variants (MAF<0.01) in *PCSK9* (N=93) and *APOB* (N=11), compared with non-carrier controls (N=9,540) from ASPREE. Results indicate that rare *APOB/PCSK9* rare variant carrier status is associated with reduced serum LDL-C and TC levels. Linear regression was used to compare statistical differences between groups adjusting for covariates. We first tested association using raw unadjusted LDL-C (**a**) and TC (**b**) levels (not adjusting for statin use), then separately using statin-adjusted *adj*LDL-C (**c**) and *adj*TC (**d**), dividing LDL-C and TC levels by 0.7 and 0.8 respectively for participants using statin medication to estimate untreated levels ^3^. Median values are shown as coloured solid lines (magenta and red). We identified statin users based on concomitant medications recorded by ASPREE at baseline. Statistical significance is denoted as *** P=<0.001. Dyslipidaemia (dotted lines) is defined as serum LDL-C higher than 160 mg/dl, or serum TC higher than 212 mg/dl ^14^. To convert values from mg/dl to mmol/l multiply by 0.02586.

For the more common *PCSK9* R46L variant (rs11591147-T, ASPREE MAF=0.03541), heterozygous and homozygous carrier status were associated with 15.5mg/dl (11.8%) lower and 25.2 mg/dl (19.2%) lower statin-corrected LDL-C levels respectively (P=<0.001) (Table 3).

## DISCUSSION

In this study, we identified rare *PCSK9* and *APOB* variants that that lower serum LDL-cholesterol levels among a population of healthy older individuals without a prior history of CHD events. Variant carrier status was associated with significantly lower serum LDL-C and TC concentrations, using statin-corrected or uncorrected levels. The prevalence of statin prescriptions in variant carriers were less than half that observed in non-carriers. Together, these results indicate that lipid-lowering genetic variants are carried by healthy elderly individuals without CHD, and play an important role in coronary disease-free survival throughout the human lifespan.

Consistent with other studies ^1-9^, we observed that loss-of-function variants in *PCSK9* and *APOB* lower serum LDL-C concentrations. However, to our knowledge, the prevalence shown in ASPREE during ageing free of atherosclerotic cardiovascular disease manifestations has not previously been demonstrated. Variants analysed were found to lower serum LDL-C and TC levels among the ASPREE population, with the difference in LDL-C concentrations between ASPREE rare variant carrier and non-carriers being ∼20 mg/dl after adjusting for statin use. At an average participant age of 75 years, this represents the effect of potentially a lifetime of exposure to genetically-determined lower LDL-C.

Meta-analysis of statin trials suggests for a 38 mg/dl reduction in LDL-C, there is a 20-22% reduction in CHD risk, in the setting of relatively short clinical trials ^27^. Mendelian randomisation studies, however, demonstrate the importance of lifetime exposure to low LDL-C, suggesting that genetically-determined low LDL-C is associated with a greater magnitude of CHD risk-reduction, compared with equivalent reduction through statin use ^28^. It is therefore likely that ASPREE rare variant carriers detected in this study, who have experienced a lifetime of exposure to genetically lower LDL-C, have benefited substantially from lower CHD risk. However, it is noteworthy that the rare variants detected likely account for only a fraction of the reduced CHD risk in the ASPREE population (N=13,131), with a range of other genetic and lifestyle factors contributing.

Strengths of the study include the sample size and unique ascertainment of the ASPREE population. The sequenced cohort was comprised of 13,131 individuals with an average age of 75 years, with no previous diagnosis of CHD or other cardiovascular events. It is rare for a population ascertained with these characteristics to be made available for genetic analysis. The sequenced cohort was the result of a unique set of circumstances made possible by the strict ASPREE inclusion criteria and age cut-off, and associated research biobank ^14,29^.

Another strength of the study is that ASPREE participants were well characterised, each receiving a medical assessment by a general practitioner at enrolment, to confirm eligibility for the trial, and to rule out previous diagnoses of CHD ^14^. This provided confidence that detected variant carriers were CHD event-free at enrolment. Other strengths of the study include the depth of sequencing, focus on canonical lipid metabolism genes with established biological effect, and stringency of variant curation used, to ensure only high-confidence variants were included in analyses.

Limitations of the study include our results not necessarily being generalizable to populations of non-European ancestry. Further, we caution the comparison of rare variant prevalence between ASPREE and reference populations such as gnomAD, due to the potential for technical artefacts introduced by differences in sequencing technologies and variant curation, and population stratification related to differences in genetic ancestry. These well-known sources of variability are compounded when attempting to compare rare variant frequencies between studies ^30^. Nonetheless, our results suggest that rare *PSCK9/APOB* variants are enriched in healthy older CHD-free individuals (Table 2), consistent with previous studies showing that LDL-C-lowering variants are associated with reduced risk for CHD and longevity ^1-9^.

Regarding the genes analysed in our study, we focused on only the most established two canonical lipid metabolism genes where rare loss-of-function variants have been demonstrated to have a high effect in reducing LDL-C levels (*PCSK9* and *APOB*) ^1,3,8,9^. We did not examine gain-of-function variants in the *LDLR* gene, or loss-of-function variants in other genes that have been associated with LDL-C reduction (e.g. *NPC1L1, LPA, APOC3, ANGPTL3/4*, and *ASGR1*). These additional genes were not included on the sequencing panel used. We also could not calculate polygenic scores due to the targeted nature of the sequencing assay.

We did not functionally validate the detected rare variants, and we were unable to estimate the degree to which each individual rare variant was associated with lower serum LDL-C levels. The relatively low number of rare variants detected necessitated combining rare variants together into a single group for blood lipid associations. A further limitation of our study was the lack of assessment of subclinical atherosclerosis in the ASPREE trial, including the absence of data on coronary artery calcium (CAC) and carotid intima-media thickness.

## CONCLUSIONS

Cholesterol-lowering *PCSK9* and *APOB* variants are carried by healthy older CHD-free individuals. Our study demonstrates the unique contribution of healthy elderly populations to exploring genetic determinants of health and lifespan. Historically, healthy elderly populations have not been the focus of large human genetic studies, mainly due to the difficulties in ascertaining large numbers of samples in this age group. However, studies focused on the healthy elderly represent an under-explored opportunity for detection of novel protective variants, especially in the context of lipid regulation and CHD ^29^. The ASPREE population, in particular, provides an exceptional platform, with a unique age and CHD-free ascertainment profile.

## Data Availability

Data are available from the corresponding author upon request.

## Acknowledgements

We thank the ASPREE trial staff in Australia and the United States, the ASPREE participants who volunteered for the trial, and the general practitioners and staff of the medical clinics who cared for the participants.

## Ethics approval and patient consent statement

Ethics approval for genetic analysis was obtained from the Alfred Hospital Human Research Ethics Committee. All participants provided informed consent for genetic analysis.

## Data availability statement

Data are available from the corresponding author upon request.

